# Is the Association Between Smoking and Depression Mediated by Inflammation? A Mendelian Randomization Study

**DOI:** 10.1101/2021.02.04.21251136

**Authors:** D. Galan, B.I. Perry, V. Warrier, D. Easton, G.M. Khandaker, G.K. Murray

## Abstract

Smoking, inflammation and depression commonly co-occur and may be mechanistically linked. However, key questions remain around the direction of association and the influence of residual confounding. We aimed to characterize the association between lifetime smoking and depression, as well as to assess the role that genetically-predicted C-reactive protein (CRP) level, an archetypal inflammatory marker, as a potential mediator for this association. We performed inverse variance weighted Mendelian randomization (MR) analyses using recently published summary-level GWAS data for lifetime smoking index, CRP levels, and depression. A subset of inflammatory-related genetic variants from the lifetime smoking GWAS were also used to assess the potential inflammatory causal pathways between smoking and depression. The analysis indicated significant reciprocal relationships between lifetime smoking and both depression (OR_Smk-Dep_ = 2.01, 95% CI 1.71-2.37, p < 0.001; O R_Dep-Smk_ = 1.09, 95% CI 1.06-1.13, p < 0.001) and CRP levels (OR_Smk-CRP_ = 1.40, 95% CI 1.21-1.55, p < 0.001; OR_CRP-Smk_ = 1.03, 95% CI 1.02-1.05, p < 0.001). These significant and positive associations were also supported by the majority of the robust MR methods performed. The reciprocal relationships between CRP levels (using >500 genetic instruments for CRP) and depression were not significant (OR_CRP-Dep_ = 1.01, 95% CI 0.99-1.04; OR_Dep-CRP_ = 1.03, 95% CI 0.99-1.07). We observed little variation in the IVW estimates between smoking and depression when we limited the genetic variants assessed to those related to inflammation or when we adjusted the analysis by CRP-levels in multivariable analysis. Our study supports potential causal associations between lifetime smoking and depression, as well as between lifetime smoking and CRP levels, but not between CRP and depression. No evidence was found that CRP mediates the relationship between smoking and depression.

## Introduction

Increasing evidence indicates a role for inflammation in the pathogenesis of mental health disorders, particularly depression^1,2^. Even though, historically, the central nervous system (CNS) has been considered an immuno-privileged region in the human body, research has shown that microglia in the CNS produce inflammatory cytokines and inflammatory processes outside of the CNS can result inflammatory responses within the CNS ^3–8^. Furthermore, diseases associated with inflammation, such as rheumatoid arthritis, diabetes mellitus, coronary heart disease, stroke have been associated with depression^9,10^.

Modifiable exposures, such as smoking behavior, have been associated with both depression and inflammation. A 2014 study in the US estimated that the smoking prevalence among participants 16% and 40% in patients without a psychiatric and those who had a diagnosis of MDD during the previous year, respectively^11^. Furthermore, observational studies seem to support a systemic elevation of serum inflammatory markers in smokers^12^, including significant increases in C-reactive protein (CRP) levels^13–15^. However, the evidence regarding direction of association and causality is unclear due to the biases inherent to observational studies^16,17^.

Mendelian randomization (MR) is an epidemiological approach that uses genetic variants as instruments to untangle the problems of reverse causation (genetic variants are fixed at conception; hence, genetically predicted levels of risk factors must precede any event) and unmeasured confounding (genetic variants show considerably less conventional confounding than phenotypic variables)^18^. If genetically-predicted values of a risk factor are associated with a specific disease outcome, then it is likely that the association between the risk factor and outcome has a causal basis^19–22^. In this manner, MR studies act like natural randomized control trials and overcome some of the biases of observational studies^23^.

Previous studies have shown through MR analyses that smoking and depression may have a bidirectional causal relationship^24^, although the mechanism through which smoking causes depression is not known. MR studies using CRP as a proxy for systemic inflammation have shown conflicting evidence regarding the association between higher CRP levels and risk for depression^20,25,26^. Recently, statistically better powered genome wide association studies (GWAS) for depression^27^ and for CRP^28^ have become available, and provide an opportunity to use improved genetic instruments, which explain a larger proportion of the variable’s variance, for causal inference to resolve prior ambiguities.

In this study, first to examine potential causality and direction of association, we conducted univariable and bidirectional MR analysis testing associations of genetically-predicted smoking behavior with CRP levels and risk of depression, and *vice versa*. Second, to examine for a potential mediating role of inflammation between smoking behavior and risk of depression, we conducted univariable MR analyses limiting the smoking exposure genetic variants to those which have been previously associated with inflammatory traits. Finally, we conducted a multivariable Mendelian randomization (MVMR) analysis to examine the associations between genetically-predicted smoking behavior and risk of depression after adjusting for genetically-predicted CRP levels using CRP levels to adjust the effect of the smoking genetic variants on depression. We used the latest GWAS data to develop statistically better powered genetic instruments compared to previous studies and used this to investigate if inflammation mediates the potential causal effect of smoking on depression.

## Methods

### Data Sources

This study used <u>publicly available summary level data</u> obtained from previously published genome-wide association studies (GWAS) for all analyses. The source and description of the GWAS summary statistics used in this report for lifetime smoking, depression, and C-reactive protein (CRP) are presented in Table 1.

**Table 1.** Characteristics of the GWAS from which the summary statistics were obtained

| Variable | GWAS Source | Population Used | Total Participants | Cases | Controls | SNPs Covered |
| --- | --- | --- | --- | --- | --- | --- |
| Lifetime Smoking | Wootton et al<sup>24</sup> | UK Biobank | 462,690 | 249,318 | 213,372 | 7,683,352 |
| Depression | PGC<sup>27,29</sup> | UK Biobank & 33 cohorts | 500,199 | 170,756 | 329,443 | 8,483,301 |
| C-Reactive Protein Levels | Han et al<sup>28</sup> | UK Biobank | 418,642 | NA | NA | 8,927,092 |

#### Lifetime Smoking Index

Lifetime smoking index was selected as the variable to represent the smoking exposure for all analyses. Wootton et al. (2019) generated this lifetime smoking index, encompassing information regarding smoking heaviness, duration, and smoking initiation and cessation. One standard deviation increase in lifetime smoking score is equivalent to an individual smoking 20 cigarettes a day for 15 years and stopping 17 years ago or an individual smoking 60 cigarettes a day for 13 years and stopping 22 years ago^24^.

#### Depression

Depression summary statistics were obtained from the Psychiatric Genetic Consortium (PGC). These data combined the meta-analysis efforts of two previously published articles^27,29^. The resulting summary statistics include a broad definition of depression, as they encompass the diagnostic criteria used as case definitions in the different cohorts included (Table 2).

**Table 2.** Depression case definitions used in the different cohorts combined for the depression GWAS summary statistics from the PGC^27,29^.

| Study | Cohort | Depression Definition |
| --- | --- | --- |
| Howard et al. (2019) | UK Biobank | (1) Self-reported past help-seeking for problems with “nerves, anxiety, tension or depression”, (2) self-reported depressive symptoms with |
|  |  | associated impairment, and (3) MDD identified from ICD-9 or ICD-10-coded hospital admission records. |
| Wray et al. (2018) | PGC29* | Structured diagnostic interviews. |
|  | deCODE | Electronic medical records. |
|  | GenScotland | Structured diagnostic interviews. |
|  | GERA | Electronic medical records. |
|  | iPSYCH | Electronic medical records. |
|  | UK Biobank<sup>+</sup> | (1) Self-reported MDD symptoms, (2) self-reported MDD treatment, or (3) electronic medical records. |
|  | 23andMe<sup>&</sup> | Self-reported diagnosis or treatment for clinical depression by a medical professional |

\* PGC29 includes 29 different cohorts<sup>30</sup>.
<sup>+</sup> UK Biobank samples were excluded to prevent overlap with Howard et al. (2019).
<sup>&</sup> 23andMe samples were excluded from the publicly available results.

#### C-reactive Protein

C-reactive protein (CRP) was selected as the proxy for systemic inflammation^6,14,15,31,32^. Han et al. (2019) generated the summary statistics used for this study from 418,642 individuals of British ancestry in the UK biobank for whom CRP levels were available and whose levels were lower than 10mg/L^28^.

### Selection of Genetic Instrumental Variable

In Mendelian randomization, for a genetic variant to be a valid IV, it must meet three assumptions: (i) the variant is associated with the exposure, (ii) the variant is not associated with any confounder of the exposure-outcome association,(iii) the variant does not affect the outcome, except possibly via its association with the exposure^33^. The process for IV selection from the GWAS summary statistics for each of the variables studied was performed following a primarily statistical approach^34^. However, in some cases, the IV selection was further refined to include biological factors, as described below.

#### Statistical selection

SNPs were considered significantly associated to the GWAS variable of interest if the GWAS p-value reported on the summary statistics was smaller than 5 × 10^−8 33^. Using multiple correlated variants representing the same effect would decrease the efficiency of the analyses and increase the risk of weak instrument bias in the estimates obtained without increasing the power of the study^35,36^. Consequently, absence of LD and independence of the final IVs selected was ascertained using the *ld_clump()* function from the *ieugwasr* R package (clumping window kb = 10000, *r*^*2*^ = 0.001, p = 0.99)^37^.

If an IV selected did not have a match in the outcome GWAS statistics, a proxy IV (in linkage disequilibrium with the original IV; *r*^2^ >0.8), was used instead. Proxy IVs were obtained using the *LDlink* R package^38^.

#### Biological selection

##### Smoking inflammatory SNPs

Out of the lifetime smoking IVs determined significant and in linkage disequilibrium, those SNPs that had been associated with inflammatory traits (p < 5 × 10^−8^), including those relating to cytokines, acute phase proteins, and immune cells, in previously published GWAS were subselected using the *Phenoscanner* R package^39^. These inflammatory-related IVs were used to further assess the role of inflammation in the association between smoking and depression.

##### CRP-cis SNPs

When assessing the role of CRP, four variants – rs1205, rs3093077, rs1130864 and rs1800947 – were selected as cis-variants, which are those located in the CRP gene region^20^. Limiting the analysis to cis-variants allows for more reliable conclusions due to the biological relevance of the variants used^34^. Clumping was performed using the *ld_clump()* function from the *ieugwasr* R package (clumping window kb = 10000, *r*^*2*^ = 0.001, p = 0.99)^37^ and the variant rs3093077 was selected as the lead CRP variant. Wald ratio MR analyses were performed to assess the effect of this variant on depression and smoking.

#### Univariable reciprocal Mendelian randomization analysis

The Inverse-variance weighted method was selected as the main method to calculate the combined effect of the selected instrumental variables in all univariable MR analyses. The IVW method is similar to a weighted regression of the effect of each specific IV on the outcome on the effect of the same IV on the exposure, restricting the intercept to zero^40,41^. All of the univariable MR analyses are represented in Figure 1 and in Table 3. The direction of the relationships was confirmed using Steiger filtering using the *steiger_filtering()* function from the *TwoSampleMR* R package^42–44^.

**Table 3.** Univariable MR analyses performed in this report. Analyses 1, 2, and 3 are the main analyses in this report and the rest are considered supplemental. Analyses 4, 5, and 6 are the reciprocal MR analysis for 1, 2, and 3, respectively. Analyses 1b, 1c, and 3b include biological factors in the IV selection.

| ID | Exposure | Outcome |
| --- | --- | --- |
| 1 | Lifetime Smoking | Depression |
| 1b | Lifetime Smoking – Inflammation | Depression |
| 1c | Lifetime Smoking – non-Inflammation | Depression |
| 2 | Lifetime Smoking | CRP |
| 3 | CRP | Depression |
| 4 | Depression | Lifetime Smoking |
| 5 | CRP | Smoking |
| 6 | Depression | CRP |

**Fig. 1.**
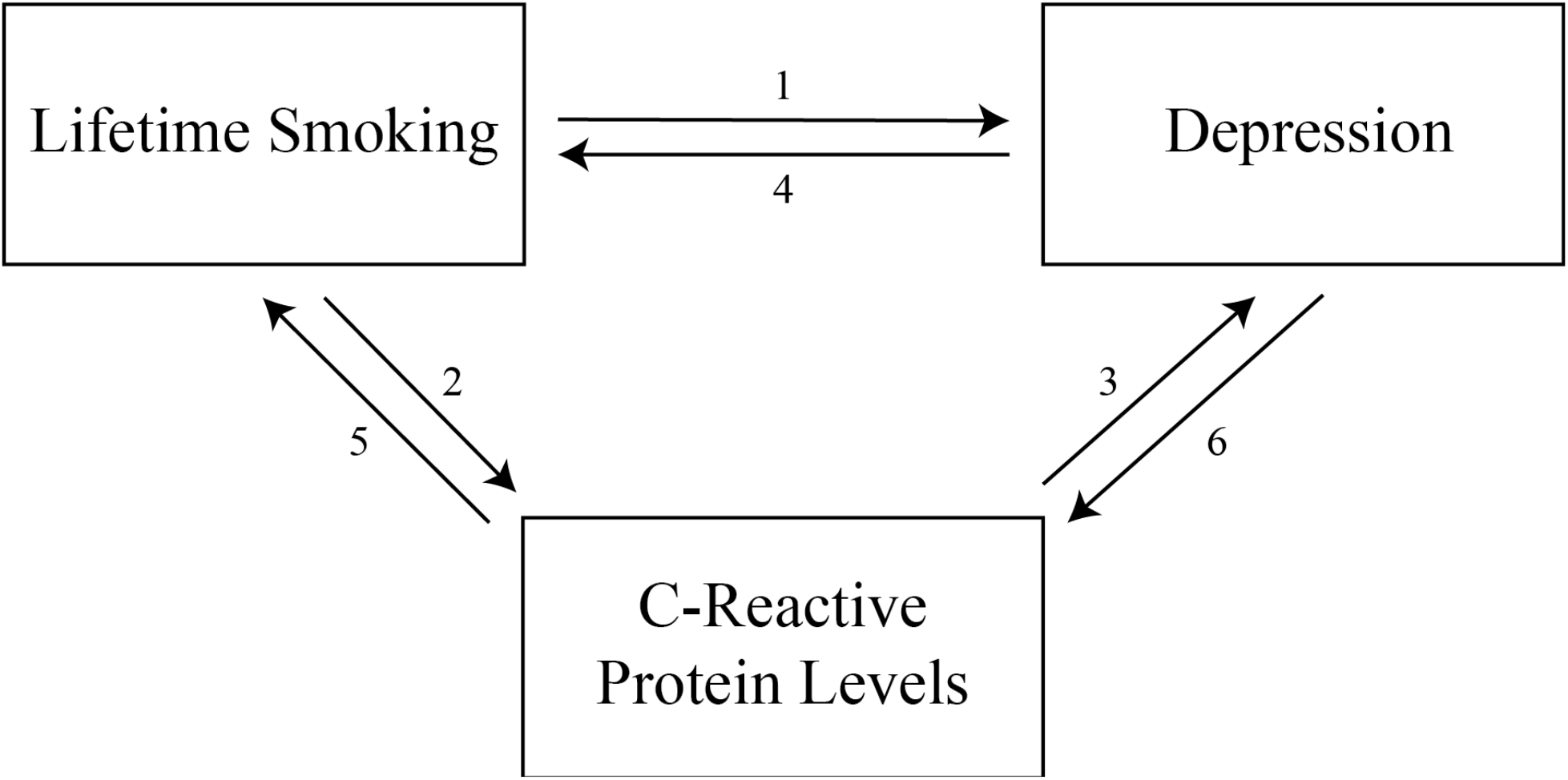
Graphic representation of the different univariable MR analyses performed. The numbers for the different analyses correspond to those listed in Table 3.

##### Heterogeneity and sensitivity analysis

For all of the analyses performed, F-statistic were calculated to assess the strength of the instruments^45^. The heterogeneity between the estimates of the different IVs used was assessed using Cochran’s Q, leave-one-out analyses, and visualizing the data with funnel, scatter, and forest plots^44,46,47^. Steiger filtering was used to assess the directionality of the relationship in the associations that were significant^42^.

The robustness of the overall estimate obtained with the IVW method was assessed using a combination of robust MR methods from different classes and working under a wide range of assumptions^42^. Four additional MR methods were used: weighted median, MR-PRESSO, MR-Egger, and contamination mixture (Table 4). Weighted median is a median based estimate that provides valid estimate even if up to 50% of the instruments are invalid. MR-PRESSO accounts for pleiotropy by detecting and removing outliers. MR-Egger accounts for pleiotropy by including an intercept term in the IVW model. Contamination mixture accounts for heterogeneity in causal mechanisms by identifying genetic instruments with similar causal estimates.

**Table 4.**
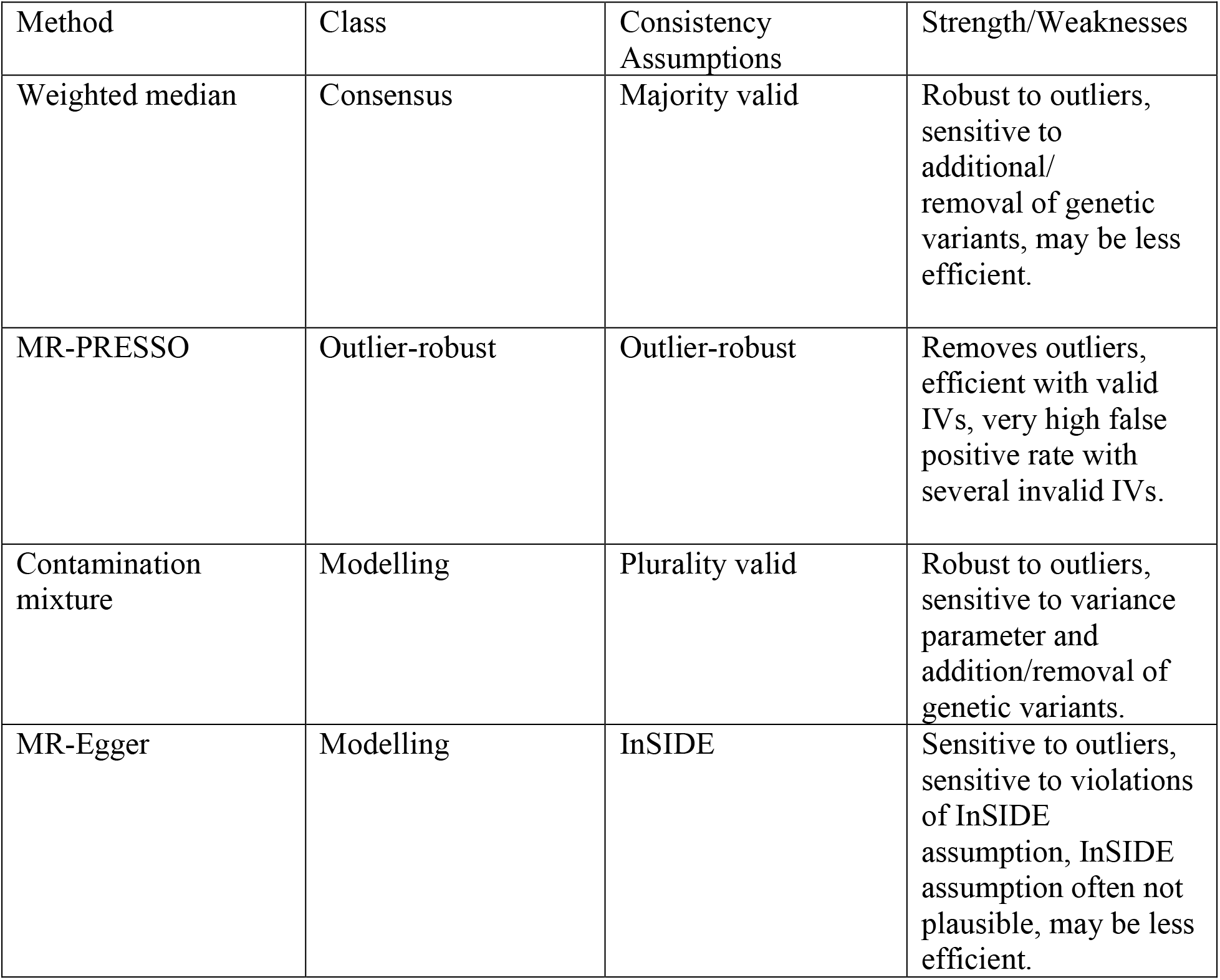
Summary comparison of the robust Mendelian randomization methods used in this report. Adapted from Slob et al. (2020).

| Method | Class | Consistency Assumptions | Strength/Weaknesses |
| --- | --- | --- | --- |
| Weighted median | Consensus | Majority valid | Robust to outliers, sensitive to additional/removal of genetic variants, may be less efficient. |
| MR-PRESSO | Outlier-robust | Outlier-robust | Removes outliers, efficient with valid IVs, very high false positive rate with several invalid IVs. |
| Contamination mixture | Modelling | Plurality valid | Robust to outliers, sensitive to variance parameter and addition/removal of genetic variants. |
| MR-Egger | Modelling | InSIDE | Sensitive to outliers, sensitive to violations of InSIDE assumption, InSIDE assumption often not plausible, may be less efficient. |

#### Multivariable Mendelian randomization analysis

(MVMR) methods allow for the estimation of the proportion of the effect of smoking directly acting on depression and the proportion potentially being mediated by CRP levels – representing systemic inflammation. IV coefficients for smoking were regressed on the SNP coefficients for depression, followed by a second regression of the coefficients of the CRP on the previously obtained coefficients^45^. The global estimate is then produced using the IVW method.

## Results

### Instrumental variable selection

The results from the IV selection process, including all of the significant SNPs in each GWAS and the final IVs selected for each variable are shown in Table 5.

**Table 5.** Instrumental variable selection using statistical methods.

| Variable | GWAS SNPs | Significant SNPs | Independents SNPs | F-statistic<sup>1</sup> |
| --- | --- | --- | --- | --- |
| Lifetime Smoking | 7,683,352 | 10,415 | 126 | 13.26 |
| CRP | 8,927,092 | 60,177 | 526 | 118.78 |
| Depression | 8,484,301 | 4,625 | 50 | 193.74 |

<sup>1</sup> F-statistic values were estimated using previously reported $R^2$ and may not represent the true value for the analyses included here.

### IVW MR Analyses Testing Association of Smoking with Depression and CRP

The IVW method supported significant associations of genetically-predicted lifetime smoking index with risk of depression (OR_Smk-Dep_ = 2.01, 95% CI : 1.71−2.37, p < 0.001), and with CRP levels CRP (OR_Smk-CRP_ = 1.40, 95% CI : 1.27−1.55, p < 0.001). The IVW method also identified a significant association between depression and lifetime smoking index (OR_Dep-Smk_ = 1.09, 95% CI : 1.06−1.13, p < 0.001), and between genetically-predicted CRP levels and lifetime smoking index (OR_CRP-Smk_ = 1.03, 95% CI : 1.02−1.05, p < 0.001) (Table 6). However, MR associations between CRP and depression were not statistically significant.

**Table 6.**
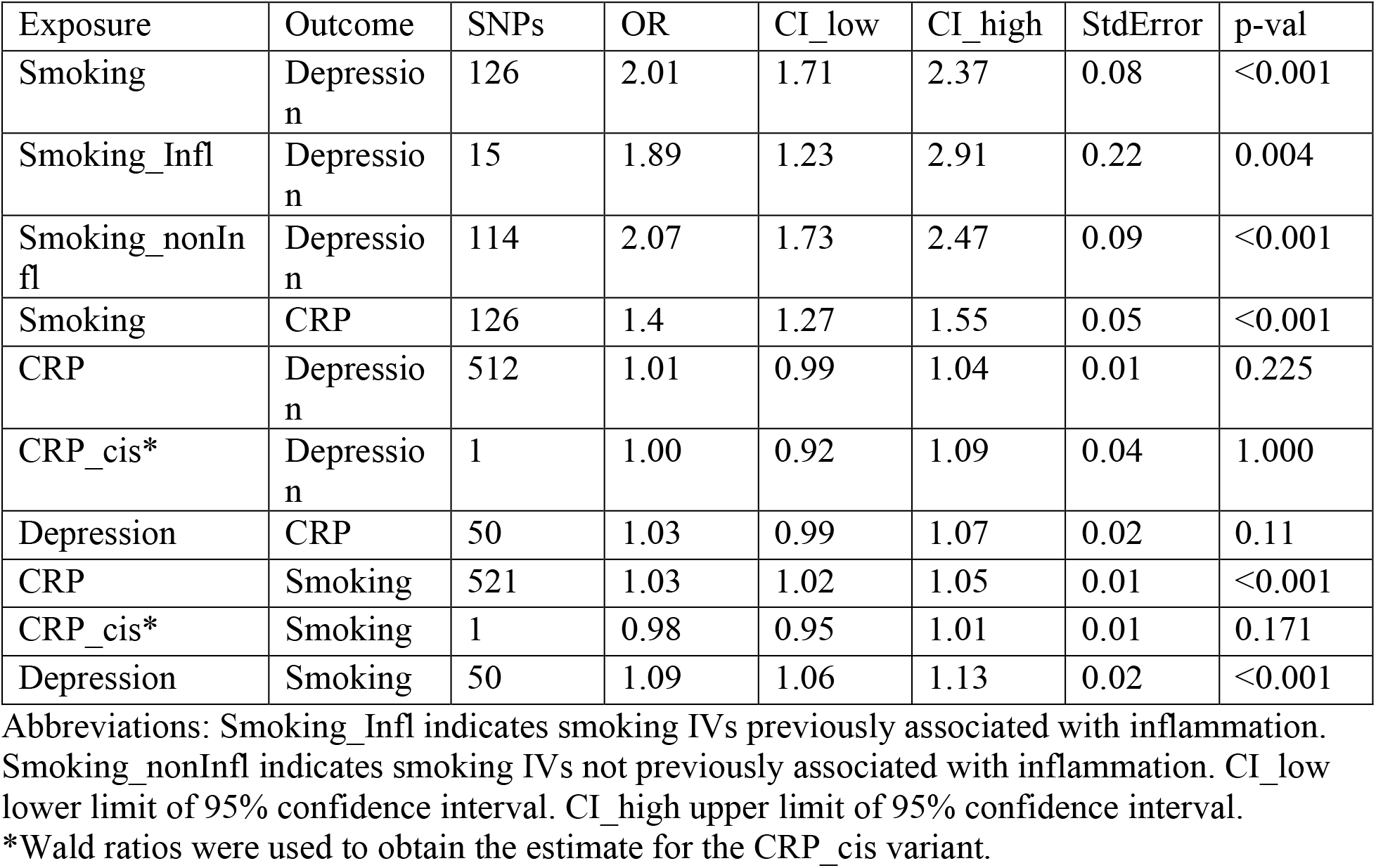
Inverse Variance Weighted Estimates for the univariable MR analyses.

### MR Analysis using inflammation-related genetic variants for smoking as IVs

Further analysis using the smoking-related genetic variants that are also associated with inflammation as IVs, the associations between smoking and depression remained significant for both inflammation-related and unrelated genetic variant sets (Table 6).

### MR Analysis using cis variants for CRP as IVs

No association was observed between the lead CRP-cis variant, rs3093077, and depression or smoking (Table 6).

#### Visual assessment of MR estimates

Visual assessment of the individual IV estimates using scatter, funnel, and forest plots indicated the presence of moderate heterogeneity among these effect estimates (Figure 2 & Supplemental Figures 1-3). However, the symmetrical distribution of individual estimates around the overall estimates suggests that the pleiotropy present is likely balanced.

**Figure 2.**
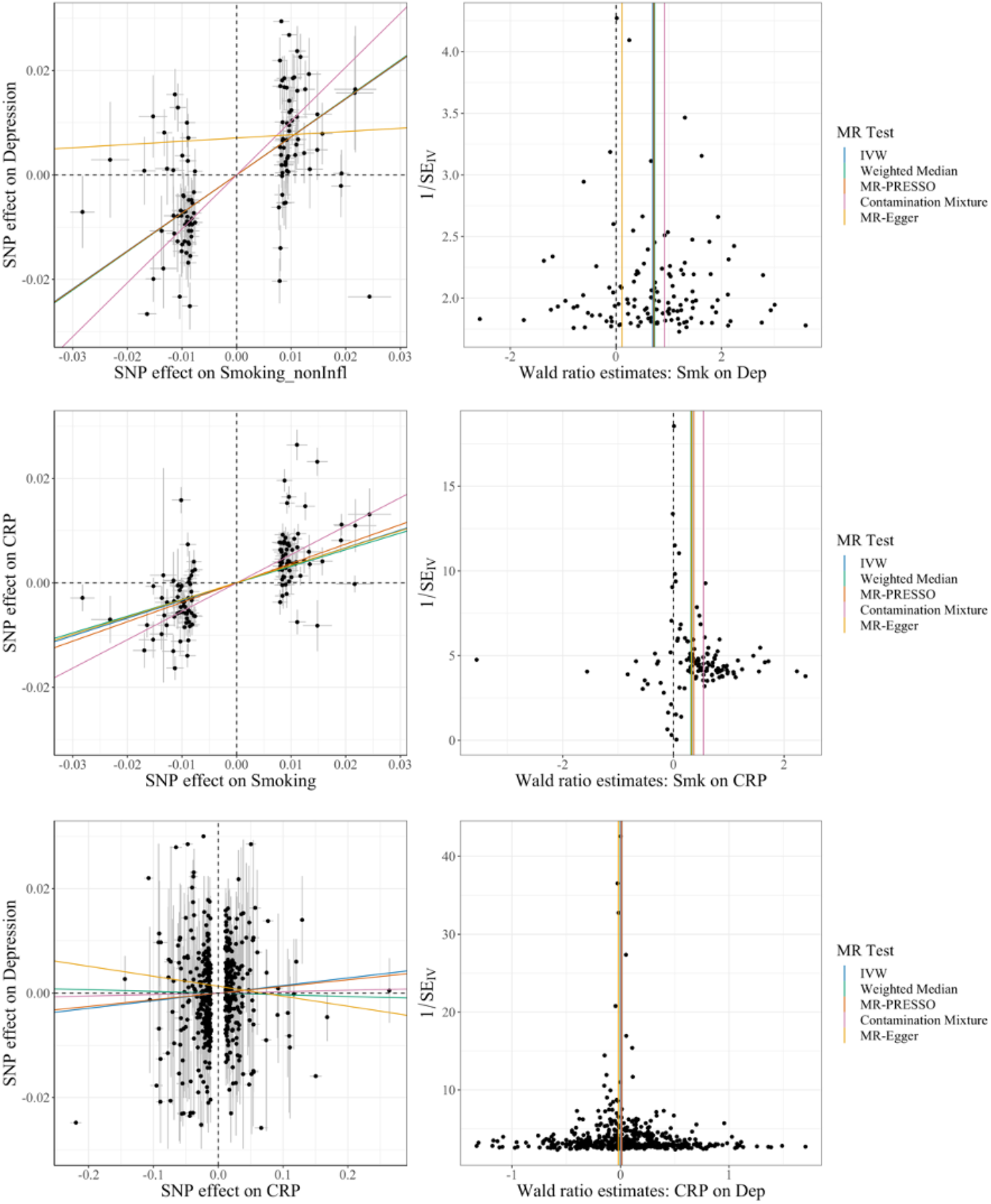
Scatter plots (left) and funnel plots (right) showing the individual IV estimates (dots) and the calculated global estimates (lines) in the main analyses.

### Results for sensitivity analyses

#### Influential IVs

Among the significant associations, every global estimate obtained through leave-one-out analyses remained significant (Supplemental Figure 2). Similarly, in non-significant associations, all of the global estimates obtained through leave-one-out analyses remained non-significant. This suggests that none of the IVs used throughout the analyses acted as an influential point.

#### Robust analyses

In the smoking-CRP, smoking-depression, and depression-smoking association, all of the global estimates calculated using robust MR methods, excluding the MR-Egger estimate, showed a similarly significant increase in the odds of the outcome and higher levels of the exposure. I^2GX^ values calculated indicate that the MR-Egger is likely to provide biased estimates; furthermore, the Q^R^ values obtained do not support a better fit of the MR-Egger data to the model when compared to the IVW model (table 9).

**Table 7.** Robust MR Estimates for the univariable MR analyses.

| Exposure | Outcome | Method | N<sub>SNP</sub><br>s | OR | StdError | 95%<br>CI_Lo<br>w | 95%<br>CI<br>Uppe<br>r | p-val |
| --- | --- | --- | --- | --- | --- | --- | --- | --- |
| Smoking | Depressio<br>n | IVW | 126 | 2.0<br>1 | 0.08 | 1.70 | 2.36 | <0.00<br>1 |
| Smoking | Depressio<br>n | Weighted<br>Median | 126 | 2.0<br>7 | 0.08 | 1.79 | 2.41 | <0.00<br>1 |
| Smoking | Depressio<br>n | MR-PRESSO | 116 | 2.0<br>5 | 0.07 | 1.79 | 2.34 | <0.00<br>1 |
| Smoking | Depressio<br>n | Contaminatio<br>n | 79 | 2.5<br>1 | NA | 2.14 | 3.13 | <0.00<br>1 |
| Smoking | Depressio<br>n | MR-EGGER | 126 | 1.1<br>2 | 0.33 | 0.59 | 2.14 | 0.727 |
| Smoking | CRP | IVW | 126 | 1.4<br>0 | 0.05 | 1.27 | 1.55 | <0.00<br>1 |
| Smoking | CRP | Weighted<br>Median | 126 | 1.3<br>8 | 0.04 | 1.28 | 1.48 | <0.00<br>1 |
| Smoking | CRP | MR-PRESSO | 111 | 1.4<br>5 | 0.03 | 1.36 | 1.54 | <0.00<br>1 |
| Smoking | CRP | Contaminatio<br>n | 87 | 1.7<br>2 | NA | 1.63 | 1.80 | <0.00<br>1 |
| Smoking | CRP | MR-EGGER | 126 | 1.3<br>9 | 0.19 | 0.96 | 2.01 | 0.08 |
| CRP | Depressio<br>n | IVW | 512 | 1.0<br>1 | 0.01 | 0.99 | 1.04 | 0.225 |
| CRP | Depressio<br>n | Weighted<br>Median | 512 | 1.0<br>0 | 0.02 | 0.97 | 1.03 | 0.838 |
| CRP | Depressio<br>n | MR-PRESSO | 504 | 1.0<br>1 | 0.01 | 0.99 | 1.03 | 0.255 |
| CRP | Depressio<br>n | Contaminatio<br>n | 380 | 1.0<br>0 | NA | 0.98 | 1.01 | 1 |
| CRP | Depressio<br>n | MR-EGGER | 512 | 0.9<br>8 | 0.02 | 0.95 | 1.01 | 0.272 |
| Depression | Smoking | IVW | 50 | 1.0<br>9 | 0.02 | 1.06 | 1.13 | <0.00<br>1 |
| Depression | Smoking | Weighted Median | 50 | 1.12 | 0.01 | 1.08 | 1.15 | <0.001 |
| Depression | Smoking | MR-PRESSO | 43 | 1.10 | 0.01 | 1.08 | 1.13 | <0.001 |
| Depression | Smoking | Contamination | 31 | 1.15 | NA | 1.13 | 1.19 | <0.001 |
| Depression | Smoking | MR-EGGER | 50 | 1.06 | 0.09 | 0.89 | 1.27 | 0.529 |
| CRP | Smoking | IVW | 521 | 1.03 | 0.01 | 1.02 | 1.04 | <0.001 |
| CRP | Smoking | Weighted Median | 521 | 1.01 | 0.01 | 1.00 | 1.02 | 0.05 |
| CRP | Smoking | MR-PRESSO | 493 | 1.03 | 0.00 | 1.02 | 1.04 | <0.001 |
| CRP | Smoking | Contamination | 349 | 1.01 | NA | 1.01 | 1.02 | 0.001 |
| CRP | Smoking | MR-EGGER | 521 | 1.00 | 0.01 | 0.98 | 1.01 | 0.533 |
| Depression | CRP | IVW | 50 | 1.03 | 0.02 | 0.99 | 1.06 | 0.11 |
| Depression | CRP | Weighted Median | 50 | 1.00 | 0.01 | 0.98 | 1.03 | 0.856 |
| Depression | CRP | MR-PRESSO | 44 | 1.04 | 0.01 | 1.01 | 1.06 | 0.009 |
| Depression | CRP | Contamination | 26 | 1.17 | NA | 1.14 | 1.20 | 0.001 |
| Depression | CRP | MR-EGGER | 50 | 1.05 | 0.10 | 0.85 | 1.28 | 0.642 |
| Smoking_Infl | Depression | IVW | 15 | 1.89 | 0.22 | 1.23 | 2.92 | 0.004 |
| Smoking_Infl | Depression | Weighted Median | 15 | 1.91 | 0.19 | 1.32 | 2.77 | 0.001 |
| Smoking_Infl | Depression | MR-PRESSO | 13 | 1.85 | 0.18 | 1.31 | 2.61 | 0.004 |
| Smoking_Infl | Depression | Contamination | 9 | 2.04 | NA | 1.34 | 3.10 | 0.012 |
| Smoking_Infl | Depression | MR-EGGER | 15 | 2.11 | 1.22 | 0.19 | 22.87 | 0.54 |
| Smoking_nonInfl | Depression | IVW | 114 | 2.07 | 0.09 | 1.73 | 2.46 | <0.001 |
| Smoking_nonInfl | Depression | Weighted Median | 114 | 2.08 | 0.08 | 1.79 | 2.44 | <0.001 |
| Smoking_nonInfl | Depression | MR-PRESSO | 103 | 2.07 | 0.07 | 1.80 | 2.39 | <0.001 |
| Smoking_nonInfl | Depression | Contamination | 72 | 2.80 | NA | 2.14 | 3.56 | <0.001 |
| Smoking_nonInfl | Depression | MR-EGGER | 114 | 1.06 | 0.34 | 0.54 | 2.10 | 0.857 |

Abbreviations: Smoking\_Infl indicates smoking IVs previously associated with inflammation. Smoking\_nonInfl indicates smoking IVs not previously associated with inflammation. CI\_low lower limit of 95% confidence interval. CI\_high upper limit of 95% confidence interval.

**Table 8.** MVMR Estimates.

|  | Exposure | Outcome | nsnp | OR | CI_Lower | CI_Upper | StdError | pval |
| --- | --- | --- | --- | --- | --- | --- | --- | --- |
| Smk-CRP-Dep | CRP | Dep | 10 | 0.92172339 | 0.70928808 | 1.19778412 | 0.133665 | 0.54198836 |
|  | Smk | Dep | 126 | 2.08138803 | 1.70563573 | 2.53991872 | 0.10158015 | 5.34E-13 |
| SmkInfl-CRP-Dep | CRP | Dep | 4 | 0.76394927 | 0.46579656 | 1.25294718 | 0.2524247 | 0.28612082 |
|  | Smk_Infl | Dep | 15 | 2.12415896 | 1.31593607 | 3.42877695 | 0.24429984 | 0.00204363 |
| SmknonInfl-CRP-Dep | CRP | Dep | 7 | 0.91312327 | 0.68136585 | 1.22370986 | 0.14937321 | 0.5428968 |
|  | Smk_nonInfl | Dep | 114 | 2.14868612 | 1.73014326 | 2.66847962 | 0.1105369 | 4.53E-12 |

Abbreviations: Smoking\_Infl indicates smoking IVs previously associated with inflammation. Smoking\_nonInfl indicates smoking IVs not previously associated with inflammation. NSNP number of SNPs.

**Table 9.** MR-Egger estimates for the main analyses and calculated I2gx and Q values.

| Exposure | Outcome | SNPs | OR | StdError | P-val | $I^2_{GX}$ | $Q'$ | $Q'$'s p-val | $Q_r$ |
| --- | --- | --- | --- | --- | --- | --- | --- | --- | --- |
| Smoking | Depression | 126 | 1.12 | 0.33 | 0.727 | 0.4 | 488.92 | <0.001 | 0.974 |
| Smoking | CRP | 126 | 1.21 | 0.33 | 0.08 | 0.98 | 1139.65 | <0.001 | 1 |
| CRP | Depression | 512 | 0.98 | 0.02 | 0.272 | 0.98 | 992.31 | <0.001 | 0.986 |

Abbreviations: Smoking_Infl indicates smoking IVs previously associated with inflammation. Smoking_nonInfl indicates smoking IVs not previously associated with inflammation. CI_low lower limit of 95% confidence interval. CI_high upper limit of 95% confidence interval.

### Multivariable MR

Using the IVW method, the MVMR analysis, using lifetime smoking as the exposure, estimated a significant OR of 2.08 for depression (95% CI : 1.71−2.54; p < 0.001), and a non-significant OR of 0.92 for CRP levels (95% CI : 0.71−1.19; p = 0.542) (Table 8). The similarity between this estimate for smoking-depression and the unadjusted one, as well as the lack of significance for the CRP term in the model, does not provide evidence to support a role for CRP as a mediator the relationship between smoking and depression.

#### Inflammation-related smoking IVs

Secondary MVMR analyses were conducted with the subset of inflammatory and non-inflammatory smoking IVs being adjusted by CRP. These two analyses provided similar estimates to the ones obtained without adjusting by CRP levels (OR_Smk−Infl_ = 2.12, 95%CI : 1.32−3.43, p=0.002; OR_Smk−nonInfl_ =2.15, 95%CI : 1.73−2.67, p<0.001) (Table 8). Again, these results do not support a role for CRP levels as a mediator for the relationship between smoking and depression.

#### Assessment of bias and reliability of MR-Egger results

As MR-Egger tends to suffer from low statistical power and is particularly susceptible to bias from weak instruments^47^, we present assessment of reliability of the MR-Egger results (Table 9). I^2^_GX_ for the data used in the different analyses (Table 9), suggested that the results from the MR-Egger analyses may be strongly biased and are not reliable; therefore, the IVW estimate is preferred^48,49^.

## Discussion

### Summary of findings

In this study, we present comprehensive Mendelian randomization analyses testing the direction and potential causality of association between smoking, CRP, and depression. Our results suggest potentially causal bi-directional associations of smoking with depression and CRP levels. However, there was no evidence for a potentially causal association between CRP levels and depression, or for a mediating role of inflammation for the association between smoking and depression. Results from multivariable Mendelian randomization analyses also suggest no evidence for the relationship between smoking and depression being mediated by CRP levels.

The results obtained were consistent through the different sensitivity and robust analyses performed. Overall, the various robust methods performed relying on different assumptions, provide strong evidence to support the potential causal role of lifetime smoking on depression.

### Smoking and depression

For the relationship between smoking and depression, our results, using improved genetic instruments compared to prior studies, are consistent with previous evidence supporting a reciprocal association between these variables. Observational evidence has shown that depression can trigger smoking commencement and make its cessation more challenging^50–54^. Moreover, smoking has been shown to lead to depression, and smoking cessation has been associated with improved depressive outcomes and decreased depression symptomatology^54–56^. Wootton et al. (2019), using IVW methods and using data partially overlapping with that used in this study, obtained an OR of 1.99 (95% CI : 1.71−2.32; p < 0.001) for the causal role of lifetime smoking on depression, and an OR of 1.10 (95% CI : 1.02−1.17; p < 0.001) for the reciprocal relationship between depression and smoking^24^. The main differences between this analysis and that in Wootton et al. (2019) is that the Howard et al (2019) depression GWAS that informed our study had a larger sample size and incorporated many cases with a broader depression definition, including broad depression (help-seeking for problems with nerves, anxiety, tension, or depression) and probable major depressive disorder. This broader definition allows for an increase in in statistical power resulting from the larger number of cases encompassed may result in a loss of specificity^57^. Nevertheless, the results obtained are very similar to those from Wootton et al. (2019) (we found an odds ratio of 2.01 for effect of smoking on depression compared to Wootton et al odds ratio of 1.99), suggesting that a despite the lack of a formal MDD diagnosis in all of the cases used, the broader depression definition used remains relevant for MR analysis^27,29^. The association between smoking and depression has important implications; it strengthens the case for primary prevention of smoking and stop-smoking initiatives, and raises the question, for future research, of whether smoking cessation initiatives are effective treatments for depression.

### Smoking and CRP

Evidence supporting a causal relationship between smoking and elevated CRP levels has been extensively documented in observational studies: with higher CRP levels in smokers^13–15,58–71^, and decreased CRP levels after smoking cessation^14,15,58,63,65–68,70,71^. This study provides new evidence to support a causal relationship between smoking behavior and CRP levels using Mendelian randomization techniques. Furthermore, this study supported a very small, but significant, effect (odds ratio of 1.03) of CRP levels on smoking lifetime index, which may be clinically negligible. Further study would be required to investigate which aspects of lifetime smoking CRP is causally associated with (as lifetime smoking is a composite measure reflecting both initiation and persistence of smoking).

### CRP and Depression

Our results did not support a potentially causal relationship between CRP and depression or *vice versa*, though several observational studies have demonstrated that patients with depression have significantly higher CRP levels when compared to those without depression^26,31,72–75^. Of previously published MR studies assessing this relationship^20,25,26^, our prior study (Khandaker et al. (2020)) found evidence for a potentially causal association between CRP levels and depression using data from the UK Biobank cohort. More recently, using the MR approach Kappelmann and colleagues have reported that inflammatory markers like CRP and IL-6 are associated with specific symptoms of depression, such as suicidality^76^. Using symptom-level data from the UK Biobank and Dutch NESDA cohorts we have reported observational and MR associations for CRP and IL-6 with somatic/neurovegetative symptoms of depression such as fatigue and sleeping difficulties^77^. Taken together, current evidence from epidemiological and genetic MR studies is consistent with inflammation being potentially causally related to certain symptoms of depression, namely somatic/neurovegetative symptoms, though MR evidence for a potentially causal role of CRP on the syndrome of depression as outcome is mixed.

### Inflammation as a mediator

The lack of difference in results of the analyses using inflammatory-related and non-inflammatory related smoking IVs, suggest that there is no clear distinct CRP mediated inflammatory causal pathway mediating the causal relationship detected between smoking and depression. Furthermore, MVMR analyses showed that adjusting the effect of smoking on depression for CRP levels did not result in a significant estimate for the effect of CRP levels.

Overall, these results do not support a role for CRP-indexed inflammation in the development of depression. This is null effect contrasts with a prior finding, using the same CRP GWAS and a similar methodology, that serum CRP is causally associated with another multifactorial etiology phenotype – namely age-related macular degeneration – indicating that the genetic instruments we used are capable of revealing positive causal relationships between serum CRP and disease. Randomized control trials have shown that using of anti-inflammatory agents, such as celecoxib or infliximab, can successfully improve depression outcomes in patients with elevated CRP levels and patients with treatment-resistant depression^78^, strongly suggesting that inflammation has a crucial role in the pathogenesis of at least certain types of depression^78,79^. If inflammation is only causally relevant in *certain subtypes* of depression with their own distinct etiology and pathogenesis, our approach would not necessarily detect this. However, our results remain consistent with the possibility that inflammation may cause depression, considered as a unitary entity, via *non-CRP mediated* processes. Our research strongly suggests that future studies examining the effect of inflammatory cytokines on depression should broaden their scope beyond CRP.

### Limitations

Lifetime smoking index and depression are both behavioral variables, with complex etiologies and hard to measure phenotypes. Furthermore, the statistical approach to IV selection use increases the risk of bias from horizontal pleiotropy^80^. However, the robust methods used, including MR-Egger, MR-PRESSO, and MR contamination mixture, allow MR analyses to be conducted in the presence of horizontal pleiotropy. The various sensitivity analyses we performed provide supporting evidence for the robustness of the findings.

It is important to note that all of the GWAS used for this study were obtained from the UK Biobank, resulting in a one-sample MR study. In one-sample MR studies, the exaggerated effect of the IVs on the exposure may lead to an overestimation of the causal effect between the exposure and the outcome assessed. This bias is called “winner’s curse” and could result in a strong bias in the obtained results^81,82^. Nevertheless, previous studies performed in non-overlapping samples have demonstrated the same direction of the effect between smoking and depression^24^. Furthermore, we used a smaller (47 independent IVs), non-overlapping CRP GWAS^83^, which showed a similar reciprocal associations between smoking and CRP, and CRP and depression (supplemental table x).

## Conclusion

The results from this study add on to the growing body of evidence supporting the bidirectionality of the causal relationship between smoking and depression. Furthermore, this study strengthens the evidence for a causal role of smoking on CRP levels. However, we do not find evidence for a potentially causal role for CRP on depression or for potentially mediating role for inflammation on the association between smoking and depression. Further research is needed to understand potential mechanisms for bidirectional association between smoking and depression.

## Data Availability

All data is publicly available from the GWAS catalog or Psychiatric Genomics Consortium website

## Author Contribution/Credit Statement

**Diego Galán**: Conceptualization, Methodology, Formal Analysis, Writing-Original Draft. **Ben Perry**: Resources, Methodology, Writing-Review & Editing. **Varun Warrier**: Conceptualization, Methodology, Writing-Review & Editing. **Golam Khandaker:** Methodology, Writing-Review & Editing. **Douglas Easton:** Writing-Review & Editing, Supervision. **Graham Murray:** Conceptualization, Methodology, Writing-Review & Editing, Supervision.

## Funding

This work was supported by a Wellcome Trust award to GMK (201486/Z/16/Z), and a National Institute for Health Research award to BIP (DRF-2018-11-ST2-018). VW is supported by the Bowring Research Fellowship (St. Catharine’s College, Cambridge) and a Wellcome Trust Collaborative Grant.

This work was supported by a Wellcome Trust award to GMK (201486 Z 16 Z) and a National Institute for Health Research award to BIP (DRF 2018 11 ST2 018). VW is supported by the Bowring Research Fellowship (St. Catharine’s College Cambridge) and a Wellcome Trust Collaborative Grant

This work was supported by a Wellcome Trust award to GMK (201486 Z 16 Z) and a National Institute for Health Research award to BIP (DRF 2018 11 ST2 018). VW is supported by the Bowring Research Fellowship (St. Catharine’s College Cambridge) and a Wellcome Trust Collaborative Grant

## Conflicts of Interest

All authors declare no conflicts of interest.

## Notes

### Competing Interest Statement

The authors have declared no competing interest.

### Author Declarations

We used publicly available data only.

